# Multi-ancestry proteogenomic analysis identifies risk proteins for intracranial aneurysms

**DOI:** 10.1101/2025.11.11.25339992

**Authors:** Chen-Yang Su, Juliano Malizia, Masashi Hasebe, Thomas Zheng, Alejandro-Mejia Garcia, Hsuan Megan Tsao, Zhaohe Lin, Ta-Yu Yang, Fumihiko Matsuda, Patrick A. Dion, Vincent Mooser, Guy Rouleau, Guillaume Butler-Laporte, Tianyuan Lu, Satoshi Yoshiji, Sirui Zhou

## Abstract

**Background:** Intracranial aneurysm (IA) and its complication, subarachnoid hemorrhage (SAH), cause morbidity and mortality, yet no preventative pharmacotherapies exist. Genome-wide association studies (GWAS) have identified risk loci for IA and SAH, but the causal proteins and pathways that connect genetic risk to aneurysm biology remain unclear.

**Methods:** We performed ancestry-stratified GWAS meta-analyses of IA and SAH in European and East Asian ancestries and linked these to circulating protein levels using proteome-wide Mendelian randomization (MR). We applied stringent instrument selection, sensitivity analyses, and colocalization, and implemented GWAS-by-subtraction to derive IA components not fully mediated by systolic blood pressure (SBP). We triangulated findings with UK Biobank observational associations, rare variant gene-burden testing in 426,295 exomes, and a French-Canadian familial IA cohort.

**Results:** Across 15,611 protein-outcome tests, 12 associations for nine proteins were significant in European ancestry. SLMAP, AMBP, ENTPD6, and PLEKHA1 were associated with increased IA risk, whereas SIRT2, JAG1, ADH4, and NAGLU were associated with decreased IA risk; ADAM23 was associated with increased SAH risk. Colocalization supported shared causal variants for ADH4, PLEKHA1, and SLMAP in IA. After removing SBP-mediated genetic effects, ADH4, JAG1, and PLEKHA1 remained associated with IA, suggesting effects not fully mediated by blood pressure. In UK Biobank, higher measured SLMAP, AMBP, and ENTPD6 levels showed concordant increases in cerebrovascular disease risk. Rare damaging JAG1 variants showed nominally higher odds of cerebrovascular disease, and a missense ENTPD6 variant was enriched in French-Canadian familial IA.

**Conclusions:** Integrating multi-ancestry genomics with large-scale proteomics implicates specific circulating proteins and pathways in IA and SAH risk. Convergent evidence prioritizes ADH4, a retinoid-pathway enzyme, and PLEKHA1, a phosphoinositide-binding adaptor in endothelial signaling, as non-SBP-mediated candidates for IA biology, with additional support for JAG1/Notch and SLMAP-related vascular pathways. These findings highlight mechanistic biomarkers and potential drug targets for aneurysm prevention that warrant experimental validation.

## Introduction

Intracranial aneurysms (IA) are cerebrovascular abnormalities caused by localized dilations of cerebral arteries due to vessel wall weakness or thinning^1^. Most intracranial aneurysms are asymptomatic at the time of detection, with a prevalence of 3% in the general population^2^, and lead to an elevated risk of subarachnoid hemorrhage (SAH) resulting from their rupture. SAH has severe consequences and is often fatal, with higher incidence reported in parts of Europe and East Asia^3,4^. Among survivors, long-term neurological morbidity is common and results in a substantial burden on healthcare systems. Despite the clinical significance of IA and SAH, there are no approved pharmacological therapies, other than targeting clinical risk factors, to halt aneurysm formation, growth, or rupture. Current curative treatment options are restricted to invasive methods^5^, specifically endovascular procedures, which involve a risk of procedure-related complications^6^. Thus, elucidating the biological mechanisms underlying aneurysm formation and rupture is critical to identify biomarkers therapeutic targets that could reduce the long-term burden of IA.

Genome-wide association studies (GWAS) have identified susceptibility loci associated with IA and aneurysmal SAH, highlighting a polygenic contribution to disease risk. Notably, a recent large GWAS meta-analysis reported 17 risk loci for IA (including both ruptured and unruptured cases)^7^. Another study further contextualized IA risk in relation to hypertension and smoking and refined the mapping of IA risk loci, underscoring shared cardiometabolic pathways and cell-type-specific programs^8^. However, the biological mechanisms linking these genetic variants to aneurysm formation and rupture remain poorly understood, and none of these discoveries have yet translated into preventative therapies. One reason may be the substantial genetic heterogeneity of IA, which complicates the identification of causal genes. Additionally, differences in IA prevalence across populations present challenges in replicating findings and generalizing risk prediction^9,10^. This gap in understanding, coupled with the absence of preventative pharmacological treatments, highlights the need for novel approaches to identify molecular biomarkers and therapeutic targets for IA and SAH.

Proteins are central to numerous biological processes, and their dysregulation can lead to disease, making them key biomarkers for diagnosis, risk prediction, and understanding disease mechanisms^11–16^. Moreover, most therapeutic drugs target proteins^17^. Recent advances in large-scale proteomic profiling offer an unprecedented opportunity to assess the causal role of proteins in disease development, identify novel biomarkers, and uncover new therapeutic targets through integrative analyses with genomic data^18–21^. GWAS of plasma proteins have identified protein quantitative trait loci (pQTLs), enabling Mendelian randomization (MR) analyses to explore the causal impact of proteins on disease^22–27^. MR uses genetic variants as proxies for lifelong differences in exposure levels, in order to infer causal effects of the exposures on disease outcomes^28,29^. Complementary strategies, such as colocalization to reduce the risk of confounding, and triangulation with observational and rare variant data, can strengthen causal inference and target prioritization.

Here, we conducted ancestry-specific GWAS meta-analyses of IA and SAH in European and East Asian populations, leveraging multiple large-scale resources, and then performed genetically stratified, proteome-wide two-sample MR using five proteomics cohorts to evaluate the effects of circulating proteins on aneurysm risk. Given that elevated blood pressure is a major causal risk factor for IA, we additionally applied GWAS-by-subtraction to partition systolic blood pressure (SBP)-related versus non-SBP genetic effects and then performed MR on the non-SBP-mediated IA component. We further prioritized signals using sensitivity analyses and colocalization, and triangulated evidence with observational associations in UK Biobank, rare variant gene burden tests in 426,295 exomes, and targeted analyses in a French-Canadian familial IA cohort. Collectively, these analyses prioritize a small set of circulating proteins as candidates for informing future mechanistic and therapeutic investigation.

## Methods

### Genome-wide association meta-analyses for IA and SAH

We performed ancestry-specific meta-analyses of GWAS of IA and SAH across European and East Asian ancestries (see **Data availability** section). Within each ancestry (European or East Asian) and outcome (IA or SAH), we conducted fixed-effects inverse-variance weighted meta-analysis in METAL^30^. The individual summary statistics contributing to meta-analyses were derived from a combination of previously published GWAS datasets, as detailed in **Supplementary Table 1**. Full details of sample sources and GWAS methodology for each contributing study are described in the original publications.

In European ancestries, the IA meta-analysis (*n* = 1,441,701; 16,594 cases; 1,425,107 controls) pooled Bakker 2020 Stage 1^7^, FinnGen non-ruptured cerebral aneurysm and SAH (release 12)^8^, and Million Veteran Program (MVP) cerebral aneurysm^28^. The SAH meta-analysis (*n* = 982,264; 10,057 cases; 972,207 controls) combined Bakker 2020 SAH^7^, FinnGen SAH (release 12)^8^, and MVP SAH^31^. In East Asian ancestries, the IA meta-analysis (*n* = 538,879; 6,418 cases; 532,461 controls) included Biobank Japan cerebral aneurysm^32^, Taiwan Precision Medicine Initiative (TPMI) intracranial hemorrhage^33,34^, China Kadoorie non-traumatic SAH^35^, and an Inuit cohort^36^. For analytical purposes, we grouped the Inuit cohort with the East Asian ancestry stratum because Inuit populations share substantial East Asian-related genetic ancestry owing to their demographic history^37,38^. The SAH meta-analysis combined Biobank Japan SAH^32^, TPMI intracranial hemorrhage^33,34^, and China Kadoorie nontraumatic SAH^35^ (*n* = 536,841; 4,447 cases; 532,394 controls).

We performed post-analysis filtering following standard filtering criteria as previously detailed^31,39^. Briefly, we applied variant-level quality control to reduce heterogeneity and frequency instability. Single nucleotide polymorphisms (SNPs) showing evidence of between-study heterogeneity were excluded (Cochran’s Q-test *P* < 0.05 or *I*^2^ > 80%). We then quantified minor allele frequency (MAF) variability per variant as MaxFreq - MinFreq and removed variants with MAF variability > 0.15 (> 15%) following standard filtering criteria^34^ and that implemented previously https://github.com/huw-morris-lab/meta-analysis. The final meta-analyses thus retained SNPs with Q-test *P* ≥ 0.05, *I*^2^ < 80%, and MAF variability ≤ 0.15.

For each of the four meta-analyses, we identified linkage disequilibrium (LD)-independent association signals using LD clumping with a clumping window of 1 Mb, *r*^2^ threshold of 0.001, and genome-wide significance level of 5 × 10^-8^. LD was computed against ancestry-matched reference panels. For European ancestry, we used a reference panel consisting of 50,000 randomly selected, unrelated individuals of European descent from the UK Biobank^40^ (UKB 50k). For East Asian ancestry, the LD structure was derived from the East Asian subset of the 1000 Genomes Project^41^ (1kGP EAS). We annotated the nearest gene for independent variants based on distance to the transcription start site.

### Proteomics study cohorts

We leveraged four large-scale proteomics studies derived from individuals of European ancestry from ARIC^18^, deCODE^19^, Fenland^20^, and UKB-PPP^21^. Details of these studies can be found in their original publications and **Supplementary Table 1**. Briefly, participants were profiled using the SomaScan v4 assay (SomaLogic) in the ARIC study comprising 4,657 circulating plasma proteins measured in up to 7,213 European American individuals, the deCODE study comprising 4,719 proteins measured in up to 35,559 Icelandic individuals, and the Fenland study comprising 4,775 proteins measured in up to 10,708 individuals. Participants in the UKB-PPP study were profiled on Olink Explore 3072 comprising 2,923 proteins measured in up to 34,557 individuals in the discovery cohort). In East Asian ancestries, we used GWAS from the East Asian subset of the UKB-PPP composed of GWAS of 2,923 proteins from 262 individuals and the Kyoto-Nagahama cohort encompassing 4,196 proteins (SomaScan v4) from 1,823 Japanese individuals.

### Genetic instrument selection

We performed LD clumping using a clumping window of 1 Mb, *r^2^* threshold of 0.001, and genome-wide significance level of 5 × 10^-8^ to identify independent variants from each of the four original studies above. Variants within 500 kb of the transcription start site of the protein-coding gene were considered *cis*-pQTLs. We used the same ancestry-matched reference panels described above (European: UKB 50k; East Asian: 1kGP EAS). Only variants with a MAF greater than 1% in these ancestry-matched panels were retained for further analysis.

### Two-sample Mendelian randomization

We estimated the effect of circulating plasma protein levels on meta-analyzed IA outcomes using proteome-wide two-sample MR in a genetically stratified manner (TwoSampleMR v.0.5.7^42^). MR relies on three instrumental-variable assumptions: (1) Relevance: the genetic instrument is associated with the exposure; (2) Independence: the instrument is independent of any confounders of the exposure-outcome relationship; and (3) Exclusion restriction: the instrument influences the outcome only through its effect on the exposure (i.e., no alternative causal pathways or horizontal pleiotropy). We excluded proteins within the major histocompatibility complex (MHC) due to the region’s complex linkage disequilibrium structure, which can confound genetic association signals and complicate interpretation of causal relationships^43^.

For European analyses, we utilized proteomic GWAS summary data from ARIC^18^, deCODE^19^, Fenland^20^, and UKB-PPP^21^ studies as exposures and assessed the effects of proteins in each cohort on European IA and SAH GWAS. For East Asian analyses, we used proteomic GWAS from the UKB-PPP^21^ study as exposures and assessed the effect of proteins meta-analyzed East Asian IA and SAH outcomes. In this context, the term "protein" refers to the aptamer (SomaScan) or antibodies (Olink) targeting the respective protein.

Summary statistics from protein GWAS (exposures) and IA outcome GWAS were harmonized using the harmonise_data() function. When a selected instrument was unavailable in the outcome dataset, we conducted a proxy SNP search using ancestry-matched LD reference panels from the instrument selection procedure. Proxy variants were identified using PLINK v.1.9^44^ with parameters --ld-window=5000, --ld-window-kb=5000, --ld-window-r2=0.8. We retained proxies with MAFs ≤ 0.42. Harmonized results are shown in **Supplementary Table 6-11**. Instrument strength was evaluated using F-statistics (**Supplementary Table 12**), with values above 10 considered indicative of robust instruments and lower likelihood of weak instrument bias^45,46^.

Causal estimates were derived using the mr() function and represent the odds ratio per 1 standard deviation increase in genetically predicted protein level. For proteins with one instrument, we applied the Wald ratio; for those with two or more instruments, we used an inverse-variance weighted random-effects model. We controlled for multiple testing by using a Benjamini-Hochberg false discovery rate (FDR)^47^ threshold of 5%, consistent with prior studies^25,48^. However, to ensure stringency, we performed the correction within each proteomics cohort and at the single-trait level. MR analyses adhered to STROBE-MR reporting guidelines^28,29^ (**Supplementary Note 1**).

### GWAS-by-subtraction

We conducted GWAS-by-subtraction^49^ to identify genetic effects on intracranial aneurysm that were not fully mediated by SBP-related pathways. European-ancestry IA summary statistics (*n* = 1,441,701; 16,594 cases; 1,425,107 controls) were obtained from the meta-analysis we conducted earlier. European-ancestry SBP GWAS summary statistics (*n* = 757,601) were taken from Evangelou et al.^50^. Subtraction analyses were performed following the GWAS-by-subtraction framework in Genomic SEM^51^, using a European ancestry subset of the 1000 Genomes Project^41^ as the LD reference panel. GWAS-by-subtraction partitions intracranial aneurysm into SBP-related effects (g*_SBP_*) and non-SBP-mediated latent genetic effects (g*_nonSBP_*) which represents IA genetic associations not fully mediated by SBP-related pathways (**see Data availability**). Genetic correlation analyses were performed using LDSC^52^.

We performed MR analyses on the g*_nonSBP_* GWAS following the same procedure described previously for the original IA GWAS and used an FDR-adjusted *P* value threshold of 5% for significance.

### Sensitivity analyses with alternative MR methods

To ensure the robustness of causal estimates, we applied multiple sensitivity checks, including heterogeneity testing, alternative MR methods (weighted median, weighted mode, MR-Egger), and Steiger directionality testing. For proteins with two or more instruments, we calculated a heterogeneity *P* value using Cochran’s Q (Q_pval) and *I^2^*. Associations with *I^2^* ≥ 0.5 and Q_pval < 0.05 were considered heterogeneous. For proteins with three or more instruments, we additionally applied weighted median, weighted mode, and MR-Egger methods. Consistency in effect direction across these methods was required. Directional pleiotropy was tested using mr_pleiotropy_test() (significant if *P* < 0.05). We also used Steiger filtering (directionality_test()) to exclude variants suggesting reverse causation.

### Colocalization

We used the colocalization method, SharePro^53^, to test whether protein levels and GWAS outcomes share causal variants for associations passing sensitivity analyses above. Analyses were conducted within 1 Mb of the lead *cis*-pQTL using default priors. Colocalization was defined as a posterior probability (PP.H4) ≥ 0.8.

### Heterogeneity analyses

To identify proteins contributing to distinct biological pathways for IA or SAH, we evaluated between-outcome heterogeneity between IA and SAH across four European cohorts: ARIC, deCODE, Fenland, and UKB-PPP. MR summary statistics were obtained per cohort, and proteins common to both outcomes were retained for comparison. For each shared protein, we conducted a fixed-effect meta-analysis using the rma() function in the metafor R package, combining MR effect estimates from IA SAH while weighting by the inverse of their variance. Heterogeneity was assessed using three statistics: (i) Cochran’s Q, which tests the null hypothesis that effect sizes are consistent across outcomes; (ii) the Q-test *P* value, where *P* < 0.05 indicates statistically significant heterogeneity; and (iii) *I^2^*, which quantifies the proportion of total variability attributable to heterogeneity rather than sampling error. Proteins showing significant heterogeneity (*P* < 0.05) were interpreted as potential candidates for differential associations with IA and SAH.

### Observational association analyses for cerebrovascular diseases

We used logistic regression to evaluate whether protein levels were associated with risk of cerebrovascular diseases in the UK Biobank^40^. Cerebrovascular disease was defined from linked hospital inpatient diagnoses (ICD-10 and ICD-9), death-registry codes, and self-reported data (**Supplementary Note 2**). After excluding participants lacking complete covariate, proteomic, or diagnostic information, 442,896 individuals remained representing 30,315 cerebrovascular disease cases and 412,581 controls. Of these participants, 39,649 had overlapping Olink proteomics data (2,975 cerebrovascular disease cases and 36,674 controls) and were analyzed. Plasma protein abundances were measured with the Olink Explore 3072 panel. For each protein, NPX values were rank-based inverse-normal transformed. Logistic regression models adjusted for age at recruitment, sex, recruitment center, Olink measurement batch, Olink sample processing time, and genetic ancestry (using the first 10 genetic principal components). We assessed the nine unique proteins which achieved FDR-adjusted *P* < 0.05 from MR analyses. Of these proteins, six were present in the UK Biobank. Thus, statistical significance was assessed at *P* < 0.05 / 6 = 8.3×10^-3^ to account for the number of unique proteins tested.

Incident associations were derived from Deng et al.’s plasma proteome atlas in 53,026 UK Biobank participants, in whom 2,920 plasma proteins were measured at baseline and linked to 660 incident ICD-10 defined diseases over a median 14.8-year follow-up^54^. We queried the public Proteome-Phenome Atlas for hazard ratios and 95% confidence intervals per standard deviation higher protein level for pre-specified cerebrovascular, aneurysmal and vascular outcomes (**Supplementary Note 3**).

### Rare variant gene-based association analysis in the UK Biobank

We leveraged whole genome sequencing data from the UK Biobank, comprising of 426,295 participants. Cerebrovascular disease cases and controls were defined as in the observational analyses. To capture the potential effects of deleterious rare variants, for each MR-prioritized gene, we constructed six annotation variant masks and applied three allele-frequency bins (singletons; MAF ≤ 0.001; MAF ≤ 0.01). Masks were:

- M1 (loss of function, LoF only): high-confidence predicted loss-of-function.
- M2 (LoF + missense 5/5): LoF plus missense called damaging by all 5 in-silico predictors.
- M3 (LoF + any deleterious missense): LoF plus missense called damaging by ≥1/5 predictors (also includes 5/5).
- M4 (LoF + all missense): LoF plus all missense.
- M5 (all coding): LoF + all missense + synonymous.
- M6 (synonymous only): negative-control mask.

Burden association tests were conducted using REGENIE^55^ using a Firth logistic regression model, adjusting for age at recruitment, sex, the first ten genetic principal components, sequencing batch, and assessment center. Analyses were restricted to individuals of European ancestry to minimize bias due to population stratification.

### Rare exonic variant analysis in a French-Canadian cohort

We analyzed exome sequencing data from a French-Canadian (FC) familial IA cohort (*n* = 234; 32 cases from six families and 202 controls) using the recruitment, sequencing, alignment, variant-calling, and quality-control procedures described in our previous publication^9^. We focused on potential FC founder variants in nine MR-prioritized genes (MAF < 0.05 in any gnomAD v2.2.1 ancestry). Two-sided Fisher’s exact test was applied to test the variant carrier status in FC IA cases compared to FC controls, using the 2×2 table and reported the odds ratio with exact 95% confidence interval (CI). We used Bonferroni correction to account for multiple testing across all 13 variants identified across the nine genes (*P* < 0.05 / 13 = 0.0038).

### Druggability assessment

We evaluated the druggability of putatively causal proteins using the druggable genome by Finan et al.^56^, DrugBank (v5.1.12)^57^, and Open Targets (v24.03)^58^. Heatmaps were constructed using the pheatmap package.

## Data availability

All contributing cohorts obtained ethical approval from their institutional ethics review boards, and all participants provided written informed consent.

- The contributing proteomics cohorts include the ARIC Study, deCODE study, Fenland study, UK Biobank, and Kyoto University Nagahama study.
- The UK Biobank has approval from the North West Multi-centre Research Ethics Committee as a Research Tissue Bank, and analyses were conducted under UK Biobank application 73958.
- Meta-analyzed IA GWAS (European and East Asian) and meta-analyzed SAH GWAS (European and East Asian) will be deposited on GWAS catalog upon publication of this study.
- The Inuit cohort included in the East Asian IA meta-analysis will be deposited on GWAS catalog upon publication of this study.
- The non-SBP-mediated IA GWAS from the GWAS-by-subtraction analysis will be deposited on GWAS catalog upon publication.

## Results

### Genome-wide association meta-analysis of intracranial aneurysm

We performed ancestry-stratified genome-wide association meta-analysis leveraging data from seven large-scale biobank and case-control resources encompassing nearly 1.5 million individuals (**Figure 1**). Analyses were conducted on IA and SAH in individuals of European and East Asian ancestry. Cohort details are provided in **Supplementary Table 1**. In European ancestries, we identified 34 and 14 LD independent variants for IA and SAH, respectively (**Supplementary Table 2** and **Supplementary Table 3**). In East Asian ancestries, we found five and two LD independent variants for IA and SAH, respectively (**Supplementary Table 4** and **Supplementary Table 5**). These two loci (*AKAP8* and *WIZ*) were novel associations for IA and SAH risk and are uniquely found in the East Asian populations.

**Figure 1.**
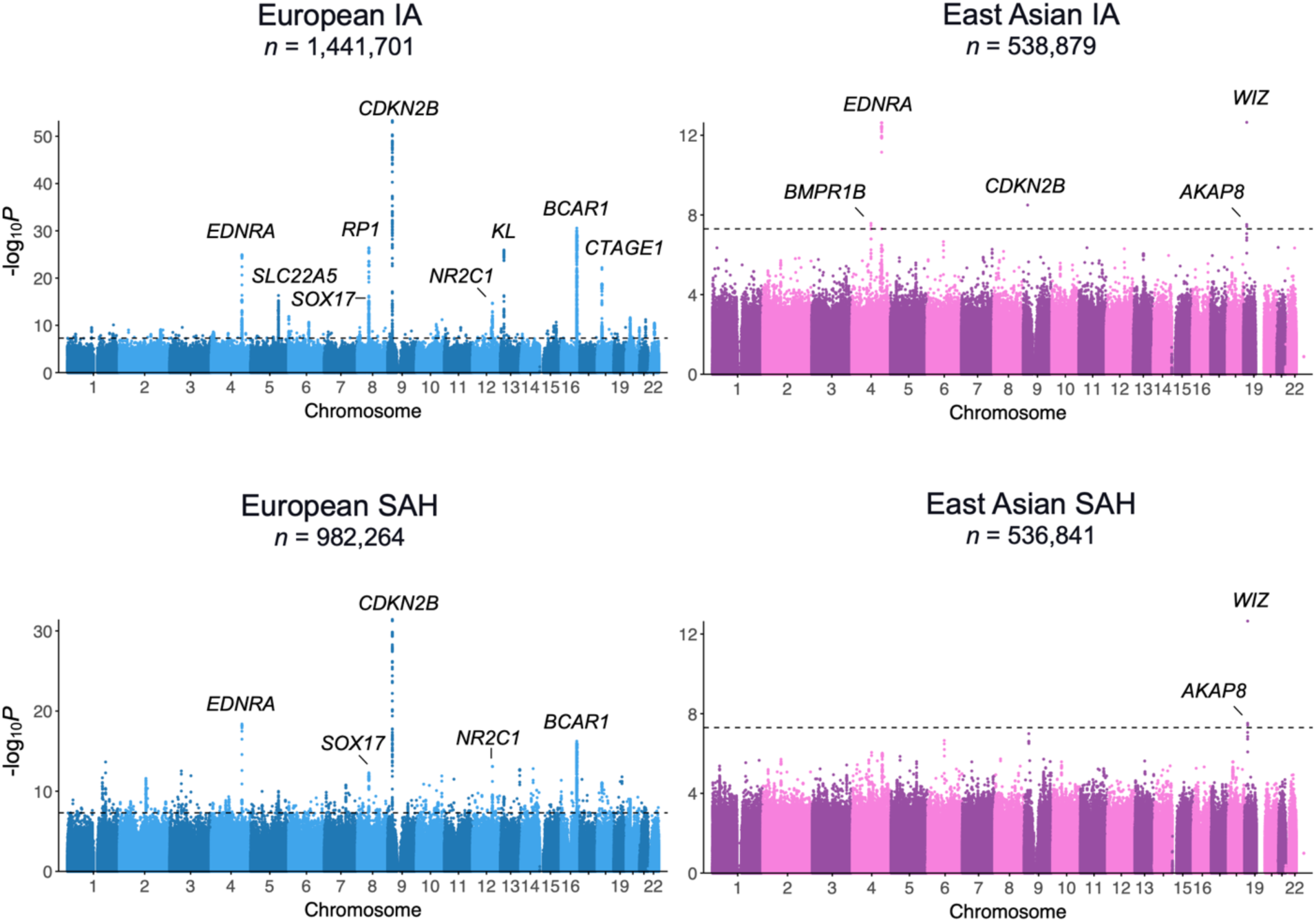
Genome-wide association results for intracranial aneurysm (IA) and subarachnoid hemorrhage (SAH) stratified by ancestry. Manhattan plots show association *P* values (-log_10_*P*; y-axis) for variants across the autosomes (chromosomes 1-22; x-axis), with alternating colors denoting adjacent chromosomes. Meta-analyses were conducted separately in European (left) and East Asian (right) cohorts for IA (top row) and SAH (bottom row). Sample sizes are given in each panel: European IA (*n* = 1,441,701); East Asian IA (*n* = 538,879); European SAH (*n* = 982,264); East Asian SAH (*n* = 536,841). The horizontal dashed line marks the genome-wide significance threshold (*P* = 5×10^-8^). Each point represents a single variant, with taller peaks indicating stronger association signals. Lead variants in linkage disequilibrium-independent regions are labeled with the nearest transcription start site gene (clumping window 1 Mb, *r^2^* < 0.001).

### Proteome-wide Mendelian randomization (MR)

Across both European (**Figure 2a**) and East Asian ancestries (**Figure 2b**), we tested 15,611 protein-disease associations in total (IA and SAH across ancestries) (**Table 1**). We confirmed that the genetic instruments explained a sufficient proportion of variance in protein levels (F > 10) across cohorts in both ancestries (**Supplementary Table 12**). To address multiple testing, we used an FDR *P* < 0.05 as the threshold for significance per cohort and identified a total of 12 FDR significant protein-disease associations (< 0.1% of the total tests). These associations were composed of 11 protein-disease pairs involving IA and a single association for SAH in European ancestries (**Figure 2c, Supplementary Figure 1,** and **Supplementary Table 13**). The 12 FDR-adjusted significant protein-disease associations in European ancestry involved nine unique proteins, with eight associated with IA and one with SAH. For IA, risk-increasing associations were observed for SLMAP, AMBP, ENTPD6, and PLEKHA1, while protective associations were seen for SIRT2, JAG1, ADH4, and NAGLU. For SAH, only ADAM23 reached significance. The direction and magnitude of effects were generally consistent across cohorts. PLEKHA1 replicated in ARIC (OR = 1.29, 95% CI 1.16–1.44, *P* = 5.54×10^-6^) and deCODE (OR = 1.22, 95% CI 1.12–1.33, *P* = 1.03×10^-5^). ADH4 replicated in Fenland (OR = 0.56, 95% CI 0.44–0.71, *P* = 1.80×10^-6^), UKB-PPP (OR = 0.62, 95% CI 0.51–0.76, *P* = 1.80×10^-6^), and ARIC (OR = 0.86, 95% CI 0.81–0.91, *P* = 1.71×10^-7^). In East Asian ancestries, no protein-disease associations were significant after FDR correction (**Supplementary Figure 1** and **Supplementary Table 14**).

**Figure 2.**
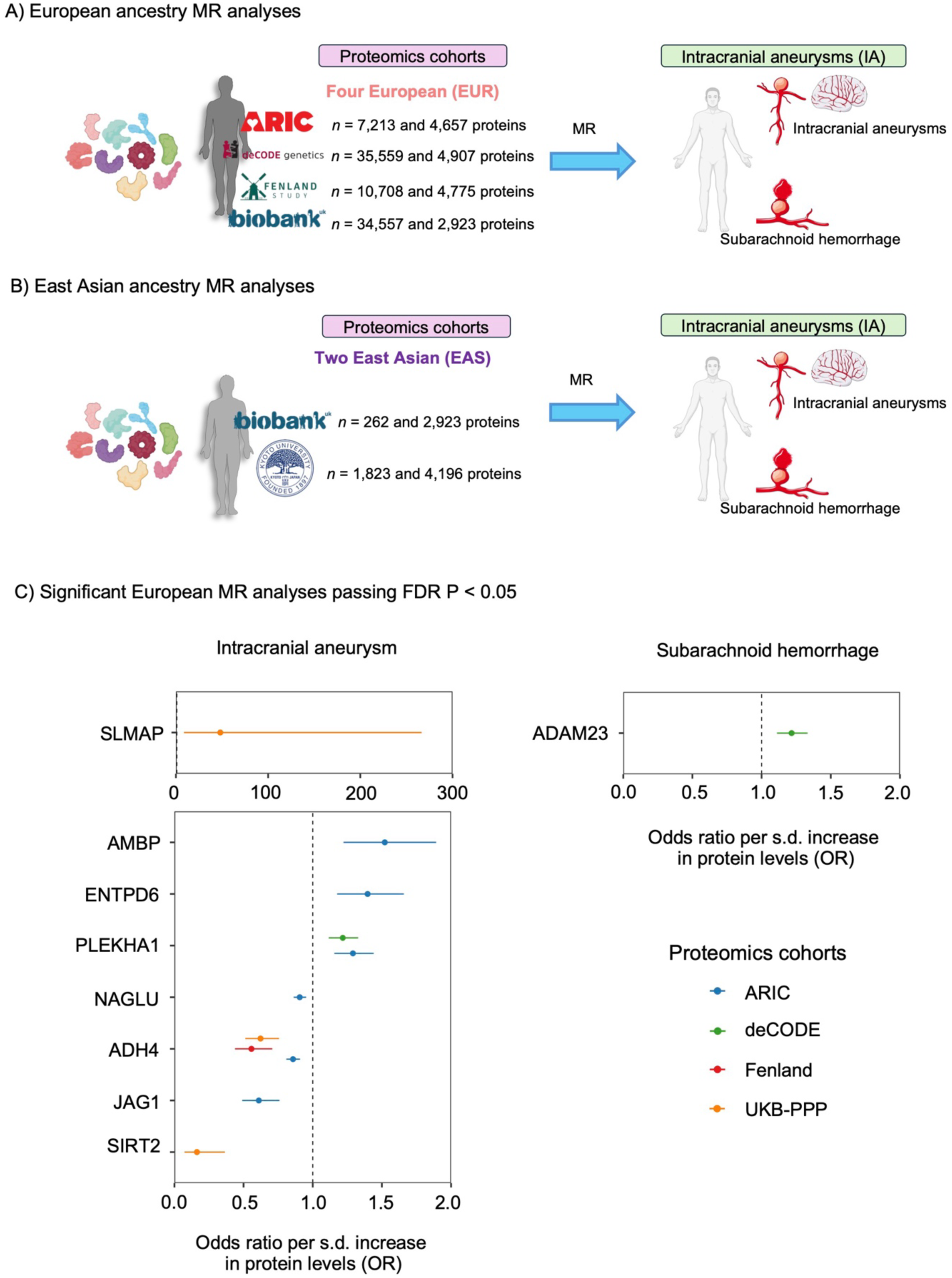
Identification of putatively causal proteins for intracranial aneurysms across ancestries. (A) European ancestry Mendelian randomization (MR) design. Two-sample MR analyses were performed using *cis*-pQTL instruments from four European proteomics cohorts: ARIC (*n* = 7,213; 4,657 proteins), deCODE (*n* = 35,559; 4,907 proteins), Fenland (*n* = 10,708; 4,775 proteins), and UKB-PPP (*n* = 34,557; 2,923 proteins) with outcomes of intracranial aneurysm (IA) and aneurysmal subarachnoid hemorrhage (SAH). (B) East Asian ancestry MR design. Conceptually identical MR analyses were carried out using instruments derived from two East Asian proteomics cohorts (UKB-PPP: *n* = 262; 2,923 proteins and Kyoto-Nagahama: *n* = 1,823; 4,196 proteins) for IA and SAH. (C) Significant European MR results (FDR-adjusted *P* < 0.05). Forest plots show odds ratios (ORs) per standard-deviation increase in genetically predicted circulating protein levels with 95% confidence intervals for IA (left) and SAH (right). Points represent cohort-specific MR estimates (colors denote proteomic cohorts: ARIC, deCODE, Fenland, UKB-PPP), and the vertical dashed line indicates the null (OR = 1). MR estimates and *P* values were obtained using the inverse-variance-weighted random-effects method when instruments included more than one variant, and the Wald ratio when the instrument comprised a single variant. Higher ORs indicate increased risk per higher protein level, whereas ORs < 1 indicate lower risk. Abbreviations: MR, Mendelian randomization; IA, intracranial aneurysm; SAH, subarachnoid hemorrhage; OR, odds ratio; FDR, false discovery rate.

**Table 1.**
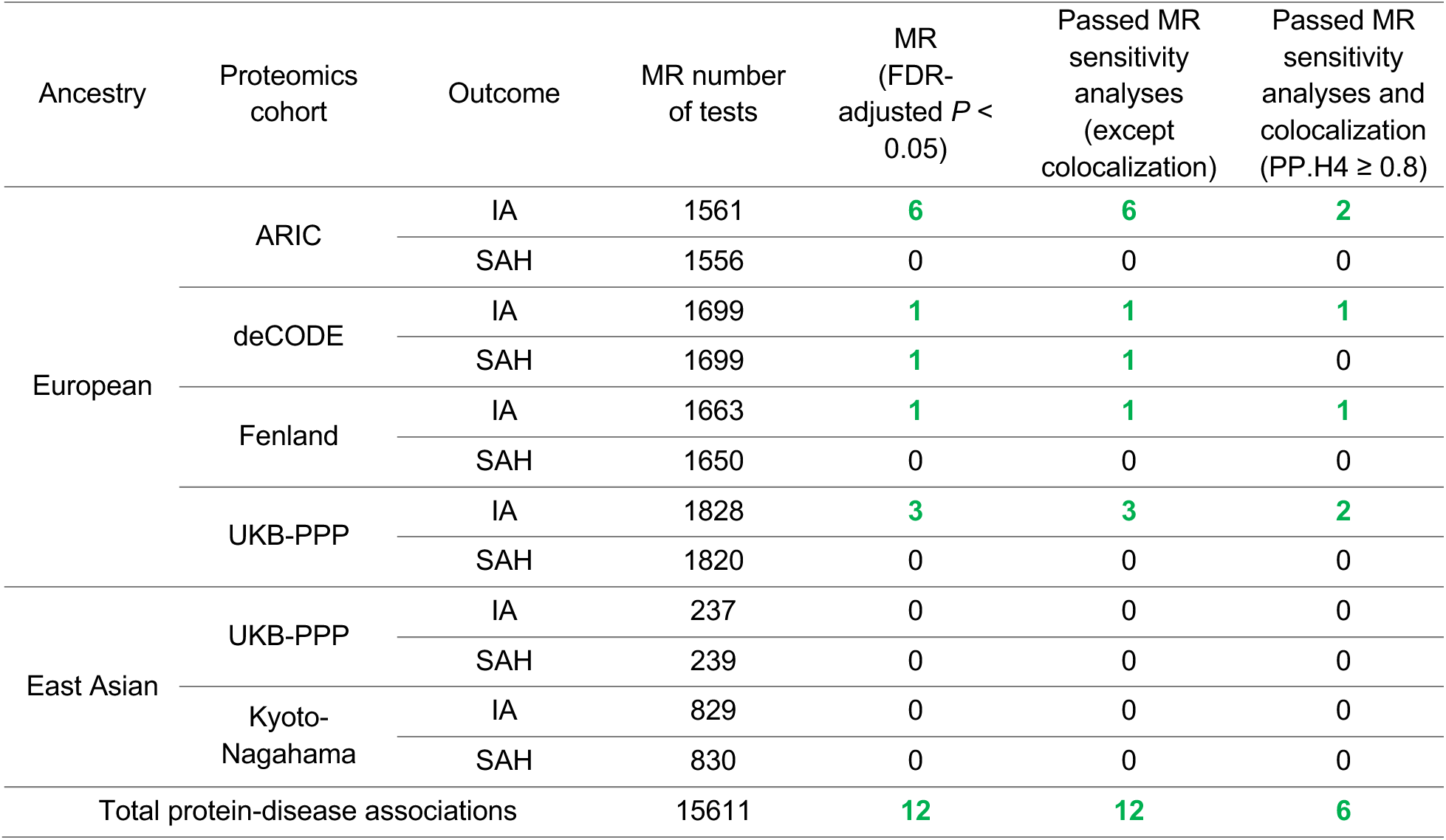
Summary of proteome-wide MR results across cohorts and ancestries. For each proteomics cohort and outcome the table reports: (i) the number of proteins with valid instruments tested (“MR number of tests”), (ii) the count reaching significance after multiple-testing correction (FDR-adjusted *P* < 0.05, controlled at the cohort-trait level), (iii) the count that also passed standard MR sensitivity checks (directionality/heterogeneity/pleiotropy; excluding colocalization), and (iv) the count additionally supported by colocalization (PP.H4 ≥ 0.8). European cohorts: ARIC, deCODE, Fenland, and UKB-PPP; East Asian cohorts: UKB-PPP and Kyoto-Nagahama. Abbreviations: MR, Mendelian randomization; IA, intracranial aneurysm; SAH, subarachnoid hemorrhage; FDR, false discovery rate; PP.H4, posterior probability of a shared causal variant.

### GWAS-by-subtraction

Given prior evidence that elevated blood pressure, a hallmark of hypertension, causally increases the risk of IA^59,60^, we used a GWAS-by-subtraction approach^49,51,61,62^ to disentangle the SBP-mediated effects of the MR-prioritized proteins in European ancestries. Upon GWAS-by-subtraction, genetic effects not mediated through SBP were less polygenic compared to genetic effects associated with SBP (**Figure 3a** and **Figure 3b**). We identified 1,640 genome-wide significant variants, 26 of which represented LD-independent loci (**Supplementary Table 15**). Additionally, the genetic correlation between SBP and IA attenuated upon removal of SBP-associated genetic signals. Prior to removal, the original meta-analyzed European ancestry IA trait demonstrated a correlation of *r_g_* = 0.256 (standard error [SE] = 0.0326). Removing SBP-associated effects using GWAS-by-subtraction attenuated the genetic correlation estimates between SBP and non-SBP IA (g*_nonSBP_*) to *r_g_* = 0.2132 (SE = 0.0324) while SBP showed high correlation with the SBP-related latent GWAS (g*_SBP_*) with *r_g_* = 0.9995 (SE = 0.0408). Taken together, these findings suggest that non-SBP-mediated genetic signals were isolated effectively.

**Figure 3.**
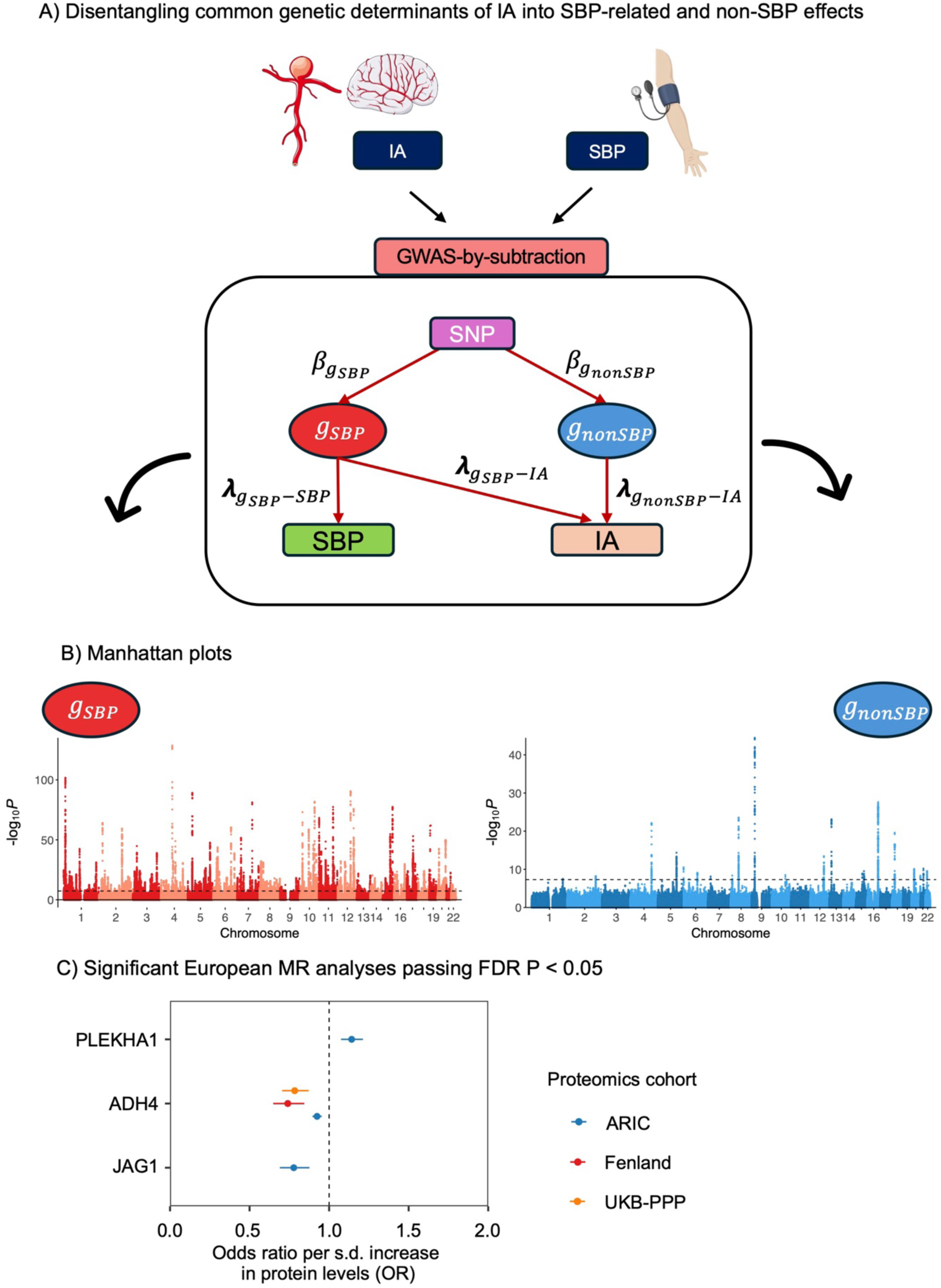
Disentangling SBP-related and non-SBP-mediated genetic effects on intracranial aneurysm and follow-up proteome-wide MR. (A) Illustration of the GWAS-by-subtraction model for systolic blood pressure (SBP)-derived effects on intracranial aneurysm (IA) in European ancestry. 𝑔*_SBP_*: SBP genetic latent factors 𝑔*_nonSBP_*: non-SBP genetic latent factors 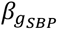: SNP effect on SBP genetic latent factor 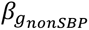: SNP effect on non-SBP genetic latent factor 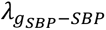: effect of SBP genetic latent factors on genetic components of SBP 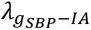: effect of SBP genetic latent factors on genetic components of IA 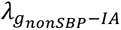: effect of non-SBP genetic latent factors on genetic components of IA (B) Manhattan plots. Genome-wide association results for 𝑔*_SBP_* (left, red) and 𝑔*_nonSBP_* (right, blue). The y-axis shows -log_10_*P* and the x-axis shows autosomes 1-22 (alternating shades). The dashed line marks the genome-wide significance threshold (*P* = 5×10^-8^). (C) Significant proteins for non-SBP-mediated IA (FDR-adjusted *P* < 0.05). Forest plots display odds ratios (ORs) per s.d. increase in genetically predicted circulating protein levels with 95% confidence intervals (CIs) for IA, using *cis*-pQTL instruments from European proteomic cohorts (ARIC, Fenland, UKB-PPP). Proteins passing FDR correction are shown (PLEKHA1, ADH4, JAG1). Points represent cohort-specific estimates, and the vertical dashed line indicates the null (OR = 1). MR estimates and *P* values were computed using the inverse-variance-weighted random-effects method when instruments contained more than one variant and the Wald ratio for single-variant instruments. Abbreviations: IA, intracranial aneurysm; SBP, systolic blood pressure; MR, Mendelian randomization; OR, odds ratio; pQTL, protein quantitative trait locus; FDR, false discovery rate.

Next, we applied proteome-wide MR to the non-SBP-mediated IA GWAS (g*_nonSBP_*) in European anceestry and found that three of the eight MR-prioritized proteins for IA had consistent effects on IA that were not acting through SBP (FDR-adjusted *P* or *q* value < 0.05; **Figure 3c** and **Supplementary Table 16**). The most consistent signal was ADH4, which showed concordant protective effects across three cohorts: ARIC (OR = 0.92, 95% CI 0.90–0.95, *q* = 1.37×10^-3^), Fenland (OR = 0.74, 0.65–0.84, *q* = 1.14×10^-2^), and UKB-PPP (OR = 0.78, 95% CI 0.70–0.87, *q* = 1.27×10^-2^). Similarly, higher JAG1 levels were protective (OR = 0.78, 95% CI 0.69–0.88, *q* = 1.89×10^-2^), whereas higher circulating PLEKHA1 levels were associated with increased IA risk (OR = 1.14, 95% CI 1.07–1.21, *q* = 1.35×10^-2^). Collectively, these results nominate ADH4 and JAG1 as putative non-SBP-mediated protective factors and PLEKHA1 as a risk-increasing factor for IA, warranting downstream validation.

### Sensitivity analyses with alternative MR methods

The 12 FDR-adjusted significant protein-disease associations passed sensitivity analyses (**Supplementary Table 17**).

### Sensitivity analyses with colocalization

To further mitigate bias from LD, we performed colocalization analyses. Of the 12 FDR-significant protein-disease associations that passed all MR sensitivity checks, six (50%) were supported by colocalization evidence, mapping to three unique proteins (**Supplementary Table 17**). For IA, ADH4 was supported by colocalization based on genetic associations in ARIC (PP.H4 = 1.00), Fenland (PP.H4 = 0.85), and UKB-PPP (PP.H4 = 0.97). PLEKHA1 was supported based on genetic associations in ARIC (PP.H4 = 0.92) and deCODE (PP.H4 = 0.93), and SLMAP was supported based on genetic associations in UKB-PPP (PP.H4 = 0.89), indicating that there was a shared causal variant underlying circulating protein levels and IA risk.

Two additional targets showed moderate colocalization with the outcomes. SIRT2 with IA (PP.H4 = 0.78) and ADAM23 with SAH (PP.H4 = 0.68). Collectively, these findings strengthen ADH4 and PLEKHA1 as putative causal mediators of IA, while suggesting that the residual MR signals involving AMBP, ENTPD6, JAG1, and NAGLU may reflect LD with nearby but distinct variants or context-dependent regulation.

### Heterogeneity analyses between IA and SAH

As an orthogonal analysis, we quantified between-outcome heterogeneity between IA and SAH MR estimates for shared proteins within each European ancestry cohort (ARIC, deCODE, Fenland, UKB-PPP). Across 6,680 IA-SAH comparisons, 58 (0.87%) showed nominal evidence of heterogeneity (Cochran’s Q *P* < 0.05), with *I*^2^ = 74.1–92.8% (**Supplementary Table 18**). The largest discordance was observed for SLMAP in UKB-PPP (*I^2^* = 92.8%; *Q* = 13.93; *P* = 1.90×10^-4^), indicating markedly different IA versus SAH effects within that cohort. In contrast, the median *I^2^* across all comparisons was 0%, consistent with low between-outcome discordance for most proteins. Together, these findings nominate a small subset of proteins with divergent associations across IA formation and rupture, suggesting partially distinct pathophysiologic pathways.

### Observational association analyses with cerebrovascular diseases

To further examine whether the MR-identified proteins contribute to organ-specific disease mechanisms, we conducted observational association analyses for cerebrovascular disease in the UK Biobank (30,315 cases and 412,581 controls) using logistic regression for the nine proteins which achieved FDR-adjusted *P* < 0.05 in the MR analyses. Six of these proteins were present in the UK Biobank and could be assessed, of which four showed significant associations in the observational analyses. Notably, results were directionally concordant between MR and observational estimates for three proteins: SLMAP (logistic regression OR = 1.09, *P* = 3.04×10^-5^), AMBP (OR = 1.26, *P* = 1.61×10^-28^), and ENTPD6 (OR = 1.07, *P* = 9.50×10^-4^), whereas ADH4 showed a discordant association (OR = 1.12, *P* = 1.27×10^-7^) (**Supplementary Table 19**).

Using Cox proportional hazards model analyses in UK Biobank from Deng et al.^54^, we next evaluated whether the six IA-prioritized proteins were associated with risk of incident cerebrovascular, aneurysmal, and vascular diseases (**Supplementary Table 20**). AMBP showed the largest and most pervasive risk elevations, particularly for coronary, peripheral arterial, hypertensive renal and aneurysmal outcomes. SLMAP and ENTPD6 displayed broadly consistent positive associations, in line with MR and logistic regression findings. ADH4, SIRT2 and ADAM23 showed more modest or selective patterns across coronary, cerebrovascular and hypertensive renal diseases. Taken together, these prospective analyses indicate that several IA-prioritized proteins—particularly AMBP, SLMAP and ENTPD6—are associated with heightened risk of a broad range of incident cerebrovascular, aneurysmal and systemic vascular diseases in UK Biobank, supporting a shared vascular aetiological component linking these proteins to intracranial aneurysm.

While JAG1, NAGLU, and PLEKHA1 were unable to be assessed, we undertook a pathway-proxy analysis in the UK Biobank by testing soluble interactors of JAG1 that were quantified in plasma as an example. Since JAG1 is a membrane-tethered ligand that is not reliably detected as a soluble analyte in the UK Biobank proteomic panels, we leveraged circulating, soluble interactors of JAG1 as pathway proxies. Among the five soluble interactors (VEGFA, DLL1, DLL4, ADAM17, and NOTCH1), only ADAM17 was not available in the UK Biobank. Logistic regression analyses suggested that higher DLL1 and VEGFA concentrations were associated with increased odds of cerebrovascular disease DLL1: OR = 1.24, *P* = 2.96×10^-25^; VEGFA: OR = 1.20, *P* = 1.67×10^-19^), both surpassing the Bonferroni threshold for four tests (α = 0.0125). By contrast, DLL4 and soluble NOTCH1 showed no evidence of association (DLL4: OR = 1.02, *P* = 0.24; NOTCH1: OR = 0.98, *P* = 0.38). These findings support a role for soluble components of the JAG1-NOTCH/angiogenic pathway in cerebrovascular disease risk and suggest complex, context-dependent regulation.

### Rare variant gene burden testing in 426,295 UK Biobank individuals for cerebrovascular disease

We analyzed whole genome sequencing data from 426,295 UK Biobank participants to perform rare variant burden tests for cerebrovascular disease in the nine MR-prioritized genes. *JAG1* showed the strongest nominal signal under the M2 mask (LoF and 5/5 deleterious missense) at MAF ≤ 0.01 and ≤ 0.001 (OR = 1.28, *P* = 0.021 for both), suggesting that carriers of rare, predicted-damaging *JAG1* variants are at increased risk of cerebrovascular disease. This is directionally concordant with MR estimates, which implied that greater JAG1 activity is protective (**Supplementary Table 21**). Additional nominal associations included *PLEKHA1* (M6 singleton, OR = 4.27, *P* = 0.041), ADH4 (M5 MAF ≤ 0.01 and ≤ 0.001, OR = 0.86, *P* = 0.052 for both associations), and *ENTPD6* (M5 MAF ≤ 0.01, OR = 1.07, *P* = 0.053). As expected for a negative control, M6 (synonymous-only) masks were broadly null, suggesting that the isolated nominal *PLEKHA1* singleton finding likely reflects noise.

### Evidence of prioritized IA genes in a French-Canadian cohort

We further assessed the contribution of potential French-Canadian (FC) founder variants in the nine MR-prioritized protein-coding genes using an FC familial IA cohort (*n* = 234; 32 cases and 202 controls). After excluding variants with MAF ≥ 0.05 in gnomAD v2.1.1, we identified 13 variants. We performed two-sided Fisher’s exact tests to test the carrier state for each variant. Among the variants evaluated, *ENTPD6* c.G67A:p.G23S (chr20:252070888, GRCh38) showed a significant case-control difference (*P* = 0.0021) (**Supplementary Table 22**). The variant was present in 8 of 32 FC IA patients (25.0%) versus 5 of 106 genotyped FC controls (4.7%). Interpreted as an enrichment in affected families, this corresponds to an odds ratio of 6.61 (95% CI 1.73–28.09), indicating substantially higher odds of carriage among affected familial IA patients. All other tested variants showed no statistically significant differences between affected and control families (two-sided *P* ≥ 0.19).

### Druggability assessment

We next assessed the druggability of the nine MR-prioritized proteins by triangulating data from the druggable genome from Finan et al.^56^, DrugBank^57^, and Open Targets^58^ (**Supplementary Table 23** and **Figure 4**). The druggable genome tiering stratified the set into one Tier 1 enzyme (ADH4), one Tier 2 enzyme (SIRT2), five Tier 3 membrane/secreted proteins (JAG1, AMBP, ADAM23, ENTPD6, NAGLU), and two unclassified targets (PLEKHA1, SLMAP). This suggests likely modality with small-molecule tractability for ADH4 and SIRT2. Concordantly, DrugBank contained bio-entity information for four of nine proteins, namely ADH4, SIRT2, NAGLU, and PLEKHA1, signaling the presence of sequence or cross-reference and, occasionally, ligand or structure information. In Open Targets, there exists no approved or clinically advanced therapeutic under development for these proteins.

**Figure 4.**
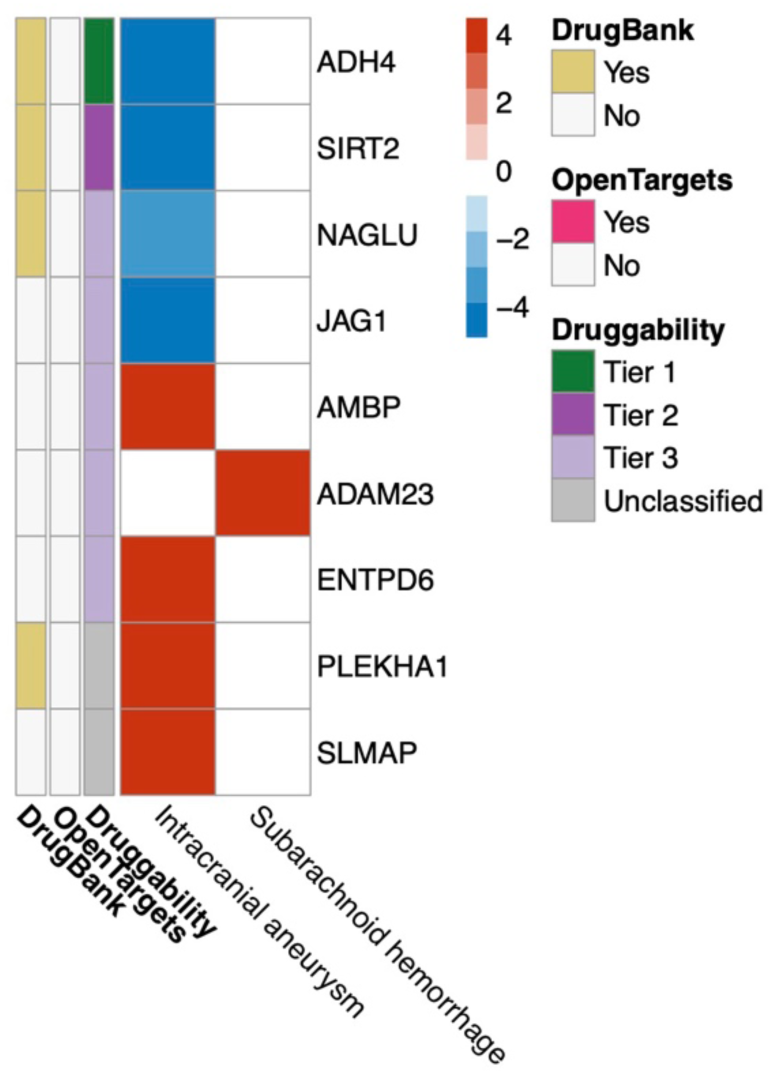
Druggability assessment of MR-prioritized proteins. Heat map summarizes proteins showing putative causal effects on intracranial aneurysm (IA) or aneurysmal subarachnoid hemorrhage (SAH) by two-sample MR (FDR-adjusted *P* < 0.05). Rows list proteins (ADH4, SIRT2, NAGLU, JAG1, AMBP, ADAM23, ENTPD6, PLEKHA1, SLMAP). Columns to the left provide target-annotation features: DrugBank (Yes/No), Open Targets (Yes/No), and druggability tier (Tier 1-3 or Unclassified). IA and SAH columns on the right display MR effects, with color indicating direction (red, risk-increasing; blue, risk-decreasing) and shade reflecting magnitude of the z score (MR estimate / standard error). MR estimates were derived using inverse-variance-weighted random-effects models for instruments with more than one variant and the Wald ratio for single-variant instruments. When a protein-disease had multiple associations, the effect estimates were averaged. Effect estimates are capped between -10 < *z* < 10 for visualization. Abbreviations: IA, intracranial aneurysm; SAH, subarachnoid hemorrhage; MR, Mendelian randomization; FDR, false discovery rate.

## Discussion

In this study, we conducted meta-analyses of IA and SAH across European and East Asian ancestries. By integrating large-scale genetic and proteomic data from five independent cohorts, we identified eight circulating proteins linked to IA risk, and one linked to SAH using MR. We further strengthened these findings through comprehensive sensitivity and colocalization analyses, investigated a GWAS-by-subtraction framework that removed genetic effects acting through blood pressure, and assessed heterogeneity between IA and SAH. We then triangulated evidence across complementary analyses and datasets, including UK Biobank observational data, rare variant burden testing, and results from a FC familial cohort. A summary of the evidence is provided in **Table 2**. These findings prioritise putatively causal proteins and pathways for mechanistic follow-up and therapeutic development.

**Table 2.** Triangulated evidence for circulating proteins implicated in intracranial aneurysm (IA) and subarachnoid hemorrhage (SAH) in European ancestries. For each protein, the table reports: (i) the number of European proteomics cohorts (of four) with a Mendelian randomization (MR) association at FDR-adjusted *P* < 0.05 and the effect direction (increasing risk vs. protective); (ii) the number of cohorts in which the MR association remained after removing genetic effects acting through systolic blood pressure (non-SBP-mediated IA from GWAS-by-subtraction); (iii) whether the signal passed MR sensitivity analyses and colocalization (PP.H4 ≥ 0.8) consistent with a shared causal variant; (iv) concordant or discordant observational associations with prevalent cerebrovascular disease in UK Biobank (logistic regression); (v) prospective associations with incident cerebrovascular, aneurysmal and other vascular diseases in UK Biobank (Cox proportional hazards models); (vi) gene based rare variant burden tests in UK Biobank exomes; and (vii) rare variant evidence in a French-Canadian familial IA cohort. Green entries indicate supportive/concordant evidence; red entries indicate discordant or negative evidence. “Nominal significance” denotes *P* < 0.05 without multiple-testing correction. Abbreviations: IA, intracranial aneurysm; SAH, subarachnoid hemorrhage; MR, Mendelian randomization; FDR, false discovery rate; SBP, systolic blood pressure; PP.H4, posterior probability that the pQTL and outcome GWAS signals colocalize (indicating a shared causal variant); LoF, loss-of-function; NA, not available since protein not measured; NS, not significant; M2 mask: LoF + missense predicted damaging by all five in-silico tools. *Results derived from Deng et al.^54^.

| Outcome | Protein | Number of cohorts with MR FDR-adjusted $P < 0.05$ (effect direction) | Number of cohorts with MR FDR-adjusted $P < 0.05$ for non-SBP-mediated IA (effect direction) | Number of cohorts passing MR sensitivity analyses and colocalization PP.H4 $\geq 0.8$ | Observational evidence for cerebrovascular disease in UK Biobank | Incident vascular disease in UK Biobank (Cox proportional hazards)* | Rare variant gene burden testing for cerebrovascular disease in UK Biobank | Rare variant analyses in French Canadian |
| --- | --- | --- | --- | --- | --- | --- | --- | --- |
| IA | SLMAP | 1 (increasing risk) | 0 | 1 | Yes, concordant with MR effect | Yes, concordant with MR effect: broad coronary, hypertensive and cerebrovascular increased risk | NS | NS |
|  | AMBP | 1 (increasing risk) | 0 | 0 | Yes, concordant with MR effect | Yes, concordant with MR effect: strong, widespread vascular and aneurysmal increased risk | NS | NS |
| | ENTPD6 | 1 (increasing risk) | 0 | 0 | Yes, concordant with MR effect | Yes, concordant with MR effect: coronary, hypertensive renal, peripheral and cerebrovascular increased risk | NS | A benign variant shows $p=0.002$ in fisher test |
|  | PLEKHA1 | 2 (increasing risk) | 1 (increasing risk) | 2 | NA | NA | Nominal significance for synonymous (negative control) | NS |
|  | NAGLU | 1 (protective) | 0 | 0 | NA | NA | NS | NS |
|  | ADH4 | 3 (protective) | 3 (protective) | 3 | No, discordant with MR effect | Inconclusive; modest, mixed pattern (partly discordant with MR) | NS | NS |
|  | JAG1 | 1 (protective) | 1 (protective) | 0 | NA | NA | Nominal significance for M2 mask | NS |
|  | SIRT2 | 1 (protective) | 0 | 0 | NS | No, discordant with MR effect: selective (ischaemic stroke, MI and arterial thrombosis) | NS | NS |
| SAH | ADAM23 | 1 (increasing risk) | Not tested | 0 | NS | Yes, concordant with MR effect: selective (hypertensive renal, peripheral and cerebrovascular) | NS | NS |

As a Tier 1 druggable enzyme, ADH4 emerged as the most robust and translationally relevant signal in our analyses. Across ARIC, Fenland, and UKB-PPP, higher genetically predicted ADH4 levels were associated with lower IA risk. These effects persisted after removing SBP-mediated components, and the pQTL signal colocalized with the IA locus in all three cohorts (PP.H4 = 1.00 for all). Interestingly, the UK Biobank observational association was directionally discordant for cerebrovascular disease, which may reflect residual confounding or differences in outcome definition. Because ADH4 is an alcohol dehydrogenase, we considered whether its association might operate via ethanol metabolism. However, class I alcohol dehydrogenases (ADH1A, ADH1B, ADH1C) are more central to hepatic ethanol oxidation at physiological concentrations, and in our MR analyses all three—ADH1A, ADH1B, and ADH1C—showed null effects on IA risk. Mechanistically, ADH4 catalyzes retinol oxidation, contributing to retinoic-acid synthesis—a pathway with vascular anti-inflammatory and matrix-stabilizing properties consistent with a protective IA effect^63,64^. In vivo, retinoic acid attenuates aneurysm progression by slowing angiotensin II-driven abdominal aortic aneurysm growth^65^, reducing vascular inflammation and matrix-metalloproteinase (MMP) activity^66^, and limiting stenosis by suppressing vascular smooth-muscle-cell migration and MMP-9^67^. Taken together, these observations suggest that the ADH4 signal is more consistent with a retinoid-centric mechanism than with ethanol clearance per se; accordingly, while reduced alcohol consumption remains a relevant public-health consideration, our genetic results point to higher ADH4, and thus greater retinoic-acid flux, as a plausible causal pathway that may stabilize the arterial wall.

PLEKHA1 showed a consistent risk-increasing association with IA, supported by significant MR estimates across two cohorts that persisted after removing SBP-related effects and demonstrated consistent colocalization. Biologically, PLEKHA1 encodes a pleckstrin-homology-domain adaptor that binds PI(3,4)P_2_ at the plasma membrane and scaffolds phosphoinositide/PI3K signaling^68,69^. This axis regulates endothelial survival, permeability, and angiogenesis^70^ which are processes central to vessel-wall remodeling in aneurysm pathophysiology^71^. Although PLEKHA1 is best known from the age-related macular degeneration locus^72^, its role within VEGF-PI3K-AKT pathways^73^ provides a coherent mechanism by which higher circulating PLEKHA1 could promote pro-angiogenic and inflammatory changes in the cerebral arterial wall.

JAG1 showed protective effects on IA risk and may act through non-SBP-mediated genetic effects. However, it colocalization evidence was equivocal. Our gene burden analysis showed nominal significance of damaging *JAG1* variants, and pathway-proxy analysis showed that soluble JAG1 interactors, including DLL1 and VEGFA, were significantly associated with cerebrovascular disease in the UK Biobank, consistent with Notch/angiogenic signaling in the vessel wall. These findings align with previous studies establishing Jagged1–Notch as essential for vascular smooth-muscle recruitment and arterial integrity^74,75^.

By contrast, SLMAP showed both colocalization based on genetic association and concordant observational association in the UK Biobank. SLMAP has been linked to endothelial dysfunction in animal resistance vessels^76^ and to human microvascular disease in association studies^77^, but not specifically to human resistance-vessel dysfunction or to increased IA risk.

We also identified extracellular matrix (ECM)-linked proteins such as AMBP, NAGLU, and ADAM23 that were prioritized through MR but did not show evidence of colocalization. Given the importance of ECM disruption in IA pathophysiology, these proteins could plausibly link circulating protein levels to arterial wall integrity. Nevertheless, further investigation is warranted to clarify the mechanisms underlying these associations.

This study has several strengths. First, we included large, ancestry-stratified GWAS meta-analyses and integrated them with large-scale proteomics datasets, enabling replication across independent cohorts, two measurement platforms (SomaScan and Olink), and two ancestral groups. Second, we conducted MR with stringent instrument selection, extensive sensitivity analyses, and colocalization to ensure causal validity. Third, we triangulated genetic with observational data and used GWAS-by-subtraction to derive IA components whose effects were not fully mediated by blood pressure alongside other orthogonal analyses to refine mechanisms and highlight biologically plausible targets.

Our study also has limitations. First, the signal specific to aneurysmal SAH was limited, despite efforts to increase power through GWAS meta-analyses. In European ancestry analyses, only one protein reached significance from MR for SAH, while no significant association was observed in East Asian ancestry analyses. Thus, conclusions regarding rupture phenotypes remain tentative and require further validation in larger GWAS. Second, the proteomic coverage, although broad, remains incomplete, and relevant proteins may not have been assayed. Third, even among the measured proteins, some lacked sufficiently strong or available genetic instruments, reducing power and precluding evaluation. Last, MR reflects lifelong, genetically proxied differences in protein levels and may not capture acute changes surrounding IA formation or rupture.

In conclusion, integrating the largest GWAS of IA across two ancestries with high-resolution population-scale plasma proteomics, we prioritized a set of proteins that appear to causally influence IA formation and rupture. These proteins provide mechanistic biomarkers that inform insights into IA pathophysiology and represent actionable entry points for future therapeutic development.

## Code availability

We used R v4.1.2 (https://www.r-project.org/), TwoSampleMR v.0.5.6 (https://mrcieu.github.io/TwoSampleMR/), SharePro v5.0 (https://github.com/zhwm/SharePro_coloc) PLINK v1.9 (http://pngu.mgh.harvard.edu/purcell/plink/), GenomicSEM (https://github.com/GenomicSEM/GenomicSEM)

## Data Availability

All contributing cohorts obtained ethical approval from their institutional ethics review boards, and all participants provided written informed consent.
- The contributing proteomics cohorts include the ARIC Study, deCODE study, Fenland study, UK Biobank, and Kyoto University Nagahama study.
- The UK Biobank has approval from the North West Multi-centre Research Ethics Committee as a Research Tissue Bank, and analyses were conducted under UK Biobank application 73958.
- Meta-analyzed IA GWAS (European and East Asian) and meta-analyzed SAH GWAS (European and East Asian) will be deposited on GWAS catalog upon publication of this study.
- The Inuit cohort included in the East Asian IA meta-analysis will be deposited on GWAS catalog upon publication of this study.
- The non-SBP-mediated IA GWAS from the GWAS-by-subtraction analysis will be deposited on GWAS catalog upon publication.

## Acknowledgments

C.-Y.S. is supported by a CIHR Canada Graduate Scholarship Doctoral Award (Funding Reference Number: 187673), an FRQS doctoral training scholarship, and a Lady Davis Institute/TD-Bank Scholarship. M.H. is supported by the Japan Student Services Organization (Graduate Scholarship for Degree-Seeking Study Abroad) and by the Watanabe Foundation (6th Toshizo Watanabe International Scholarship). The funders had no role in the study design, data collection and analysis, decision to publish, or preparation of the manuscript.

## Author contributions

Conception and design: C.-Y.S., S.Z.

Methodology: C.-Y.S., S.Z.

Data curation: C.-Y.S., J.M., C.L., S.Z.

Data Analysis: C.-Y.S.

Visualization: C.-Y.S.

Writing—Original Draft: C.-Y.S.

Writing—Review and Editing: All authors

Supervision: S.Y., S.Z.

Project administration: S.Z.

Funding acquisition: S.Z.

---

**Supplementary Figure 1.**
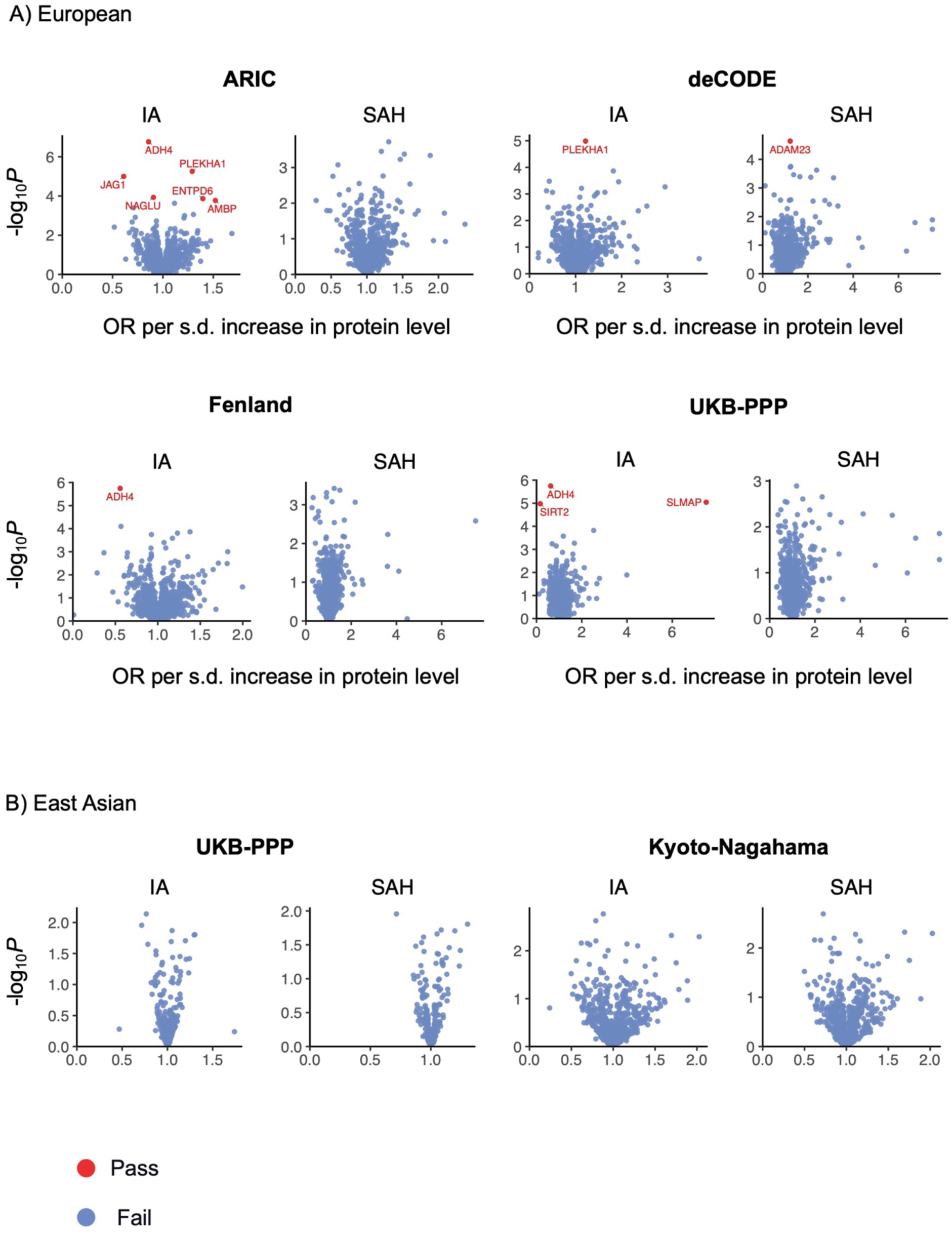
Cohort-specific volcano plots for proteome-wide MR across ancestries. (A) European cohorts (ARIC, deCODE, Fenland, UKB-PPP) and (B) East Asian cohorts (UKB-PPP, Kyoto-Nagahama). Each panel shows the MR odds ratio (OR) per s.d. increase in genetically predicted circulating protein level (x-axis) versus -log_10_*P* (y-axis) for intracranial aneurysm (IA) and aneurysmal subarachnoid hemorrhage (SAH). Points represent individual proteins; red points (“Pass”) meet the multiple-testing threshold (FDR-adjusted *P* < 0.05 within the cohort-outcome analysis) and are labeled by protein-coding gene; blue points (“Fail”) do not. OR > 1 indicates higher risk per higher protein level; OR < 1 indicates lower risk. MR estimates and *P* values were derived using inverse-variance-weighted random-effects models for instruments with more than one variant and the Wald ratio for single-variant instruments. Abbreviations: MR, Mendelian randomization; IA, intracranial aneurysm; SAH, subarachnoid hemorrhage; OR, odds ratio; FDR, false discovery rate.

**Supplementary Note 1:**
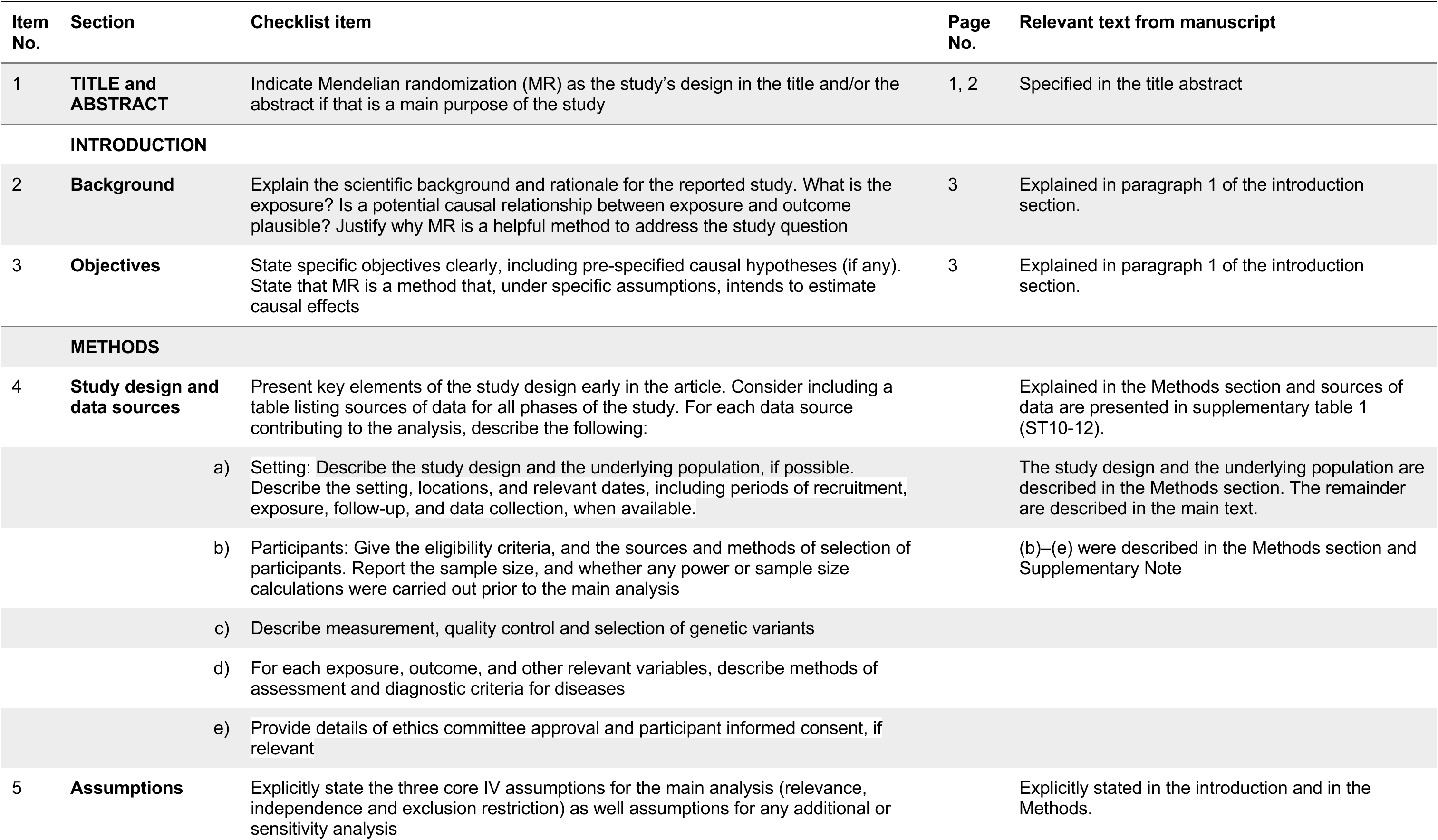
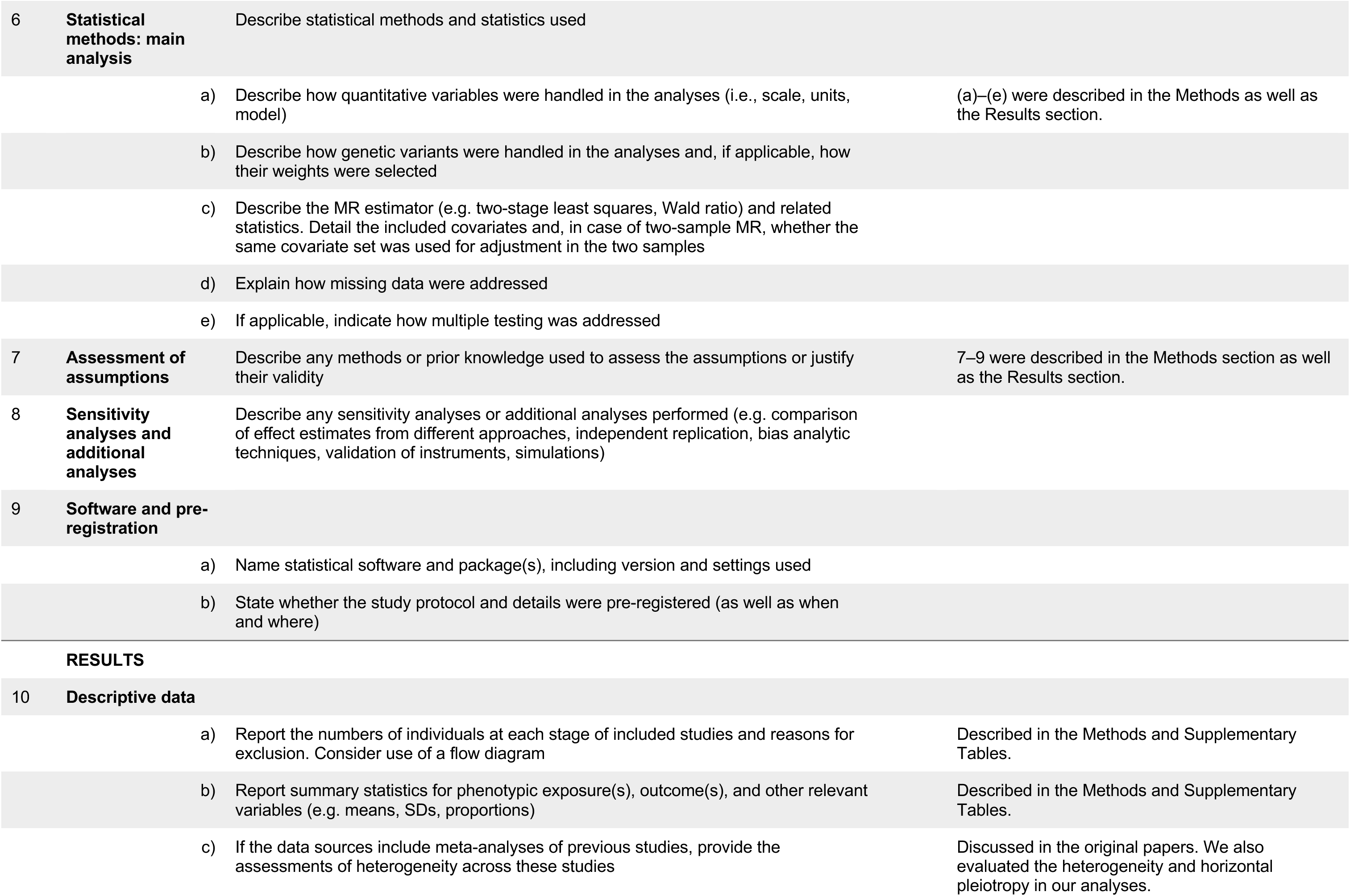
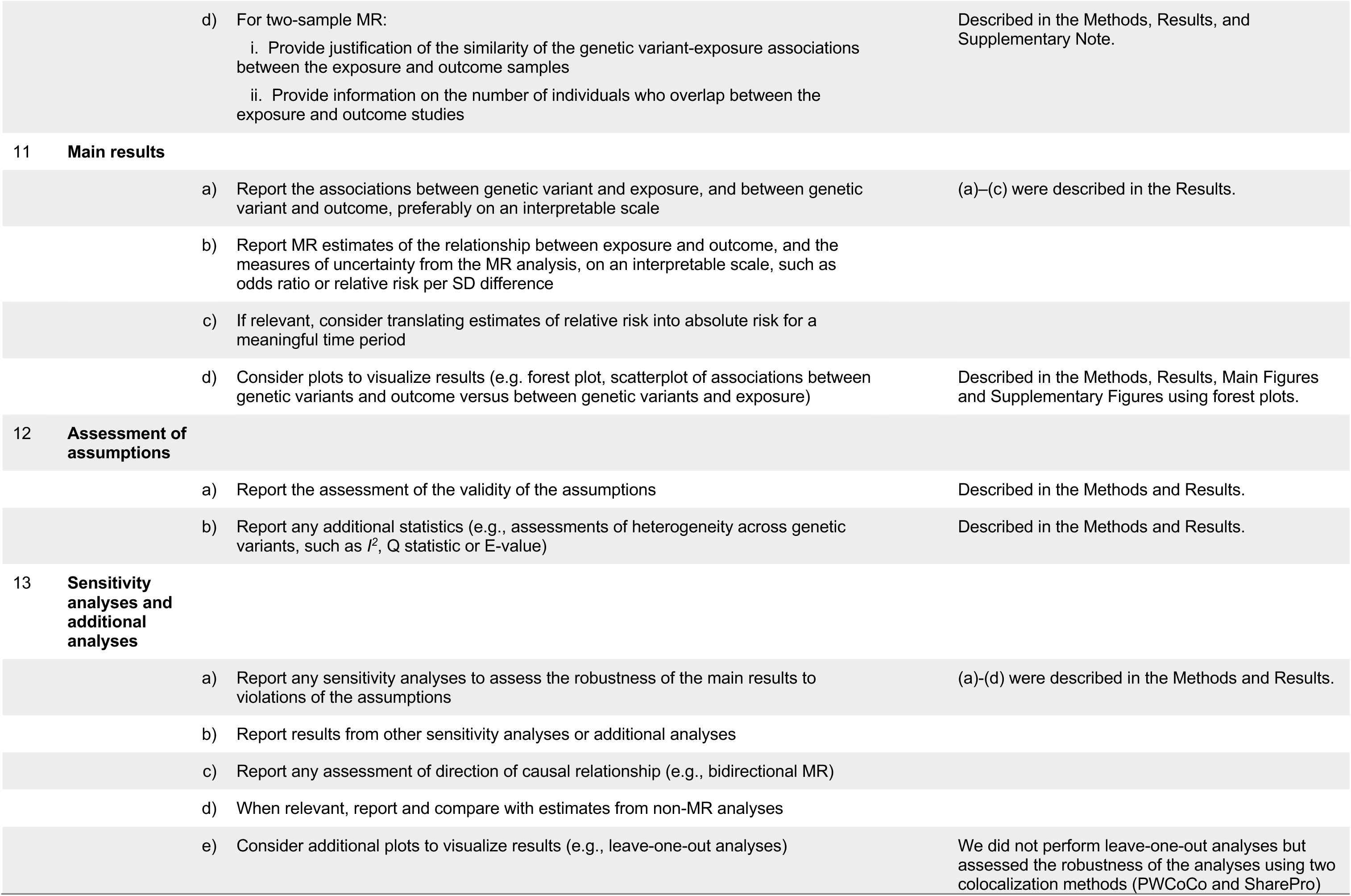
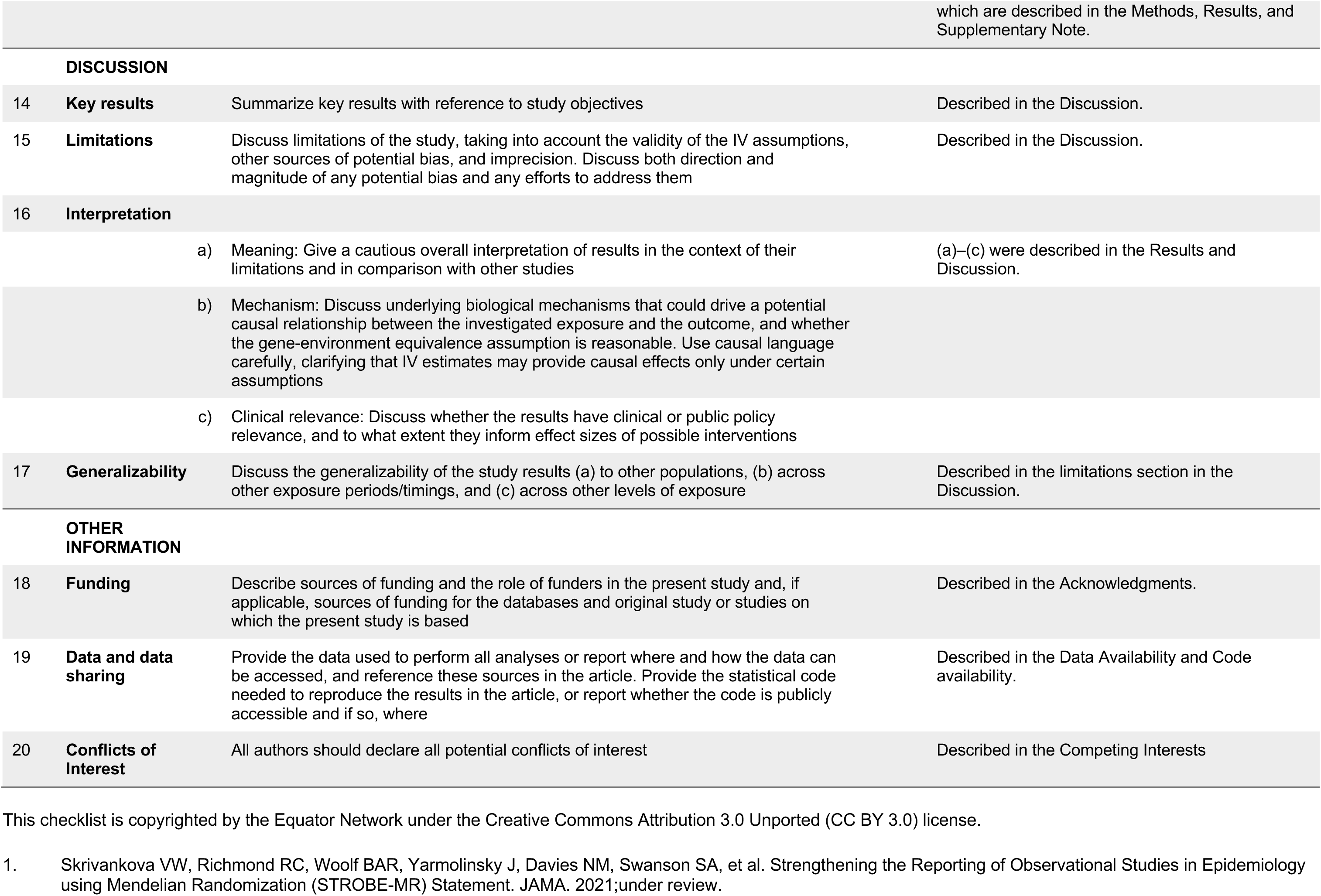
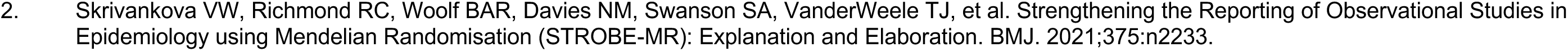
STROBE-MR checklist of recommended items to address in reports of Mendelian randomization studies^1 2^. Note: Page number will be added at the proof-reading stage.

**Supplementary Note 2.**
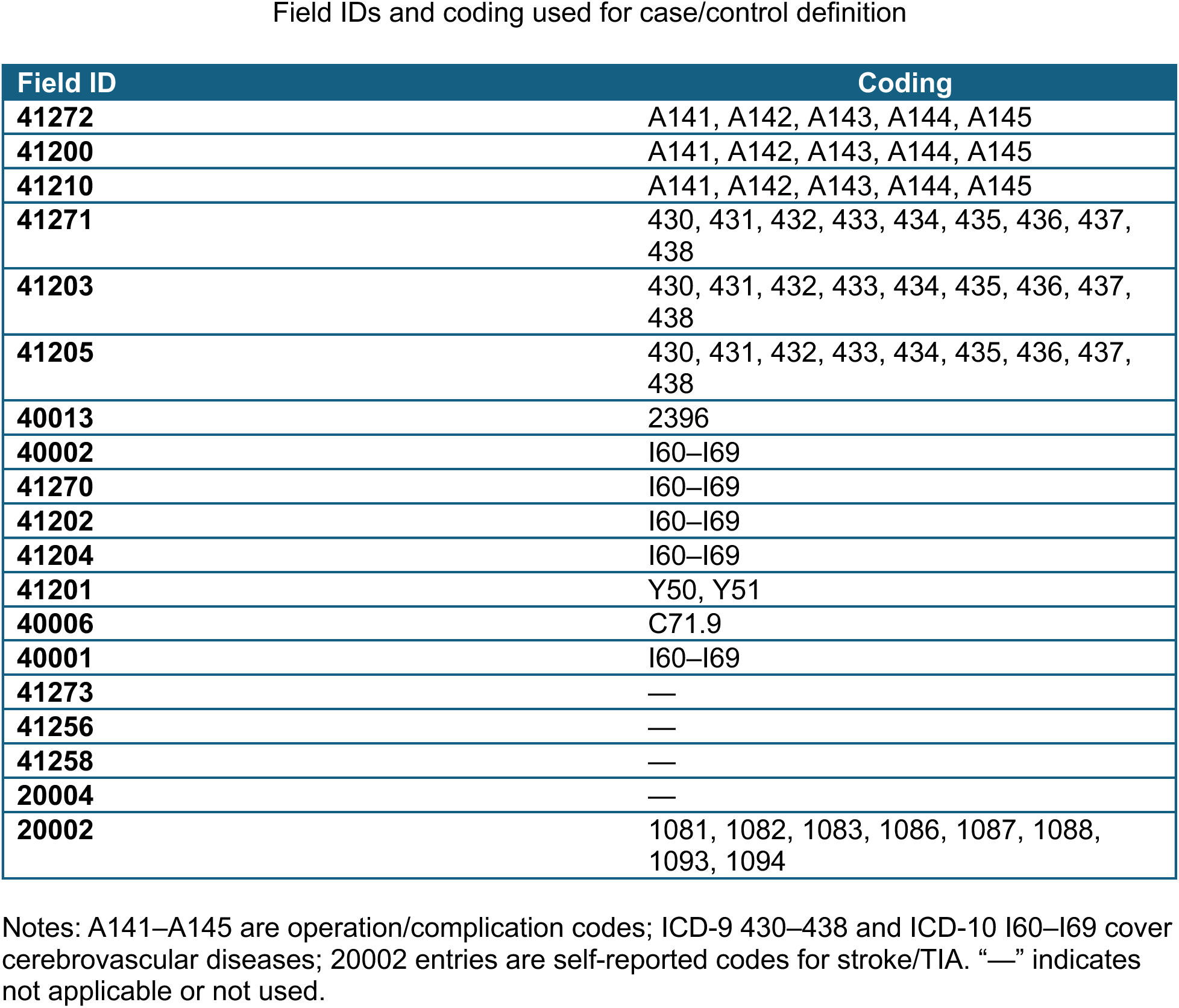
Cerebrovascular disease case definition codes in the UK Biobank. Field IDs and coding used for case/control definition

## Supplementary Note 3. Incident vascular disease phenotypes considered in UK Biobank proteome-phenome analyses

For the Cox proportional hazards analyses, we queried Deng et al.’s Proteome-Phenome Atlas for all incident endpoints available for the six IA-prioritised proteins. Among the 660 incident ICD-10-defined diseases in the atlas, 61 unique vascular and cerebrovascular phenotypes were returned at least once. We then pre-classified these into two tiers based on their relevance to intracranial aneurysm and cerebrovascular pathology.

### Tier 1 (core cerebrovascular/arterial phenotypes)

Direct cerebrovascular or arterial disease outcomes and aneurysm phenotypes: *Stroke, including SAH; Stroke, excluding SAH; Ischaemic stroke, excluding all haemorrhages; Transient ischaemic attack; Sequelae of cerebrovascular disease; Other specified cerebrovascular diseases, other cerebrovascular disorders in diseases classified elsewhere; Cerebral atherosclerosis; Aortic aneurysm; Abdominal aortic aneurysm (AAA); Other aneurysm; Peripheral artery disease; Other peripheral vascular diseases; Other diseases of arteries and capillaries*.

### Tier 2 (upstream drivers / systemic atherosclerotic phenotypes)

Vascular risk factor and systemic atherosclerotic outcomes that are biologically adjacent to IA and cerebrovascular disease and were used to provide broader vascular context: *Hypertension; Hypertension, essential; Hypertensive renal disease; Atherosclerosis, excluding cerebral and coronary sclerosis; Ischaemic heart disease, wide definition; Coronary atherosclerosis; Angina pectoris; Unstable angina pectoris; Major coronary heart disease event; Myocardial infarction, strict; Myocardial infarction, with ST-elevation; Myocardial infarction, without ST-elevation; Arterial embolism and thrombosis of lower extremity artery; Other arterial embolism and thrombosis*.

Incident hazard ratios and 95% confidence intervals for these Tier 1-2 outcomes were extracted for each of the six IA-prioritised proteins and summarised in **Table 2** and **Supplementary Table 20**.

